# Gender Based Differences in Risks and Comorbidities in Patients Hospitalized with Acute Exacerbation of COPD: A Retrospective Observational study in Eastern-Nepal

**DOI:** 10.1101/2021.07.14.21260238

**Authors:** Nischit Baral, Nabin R. Karki, Prem Raj Parajuli, Laura Bell, Bidhan Raj Paudel, Anish C. Paudel, Nirajan Adhikari, Pankaj Luitel, Narendra Bhatta

## Abstract

**Background:** Acute Exacerbation of Chronic Obstructive Pulmonary Disease (AECOPD) share a complex relationship with gender, risk, and co-morbidities. There is paucity of data on the gender-based differences in the prevalence of risks and co-morbidities in AECOPD in Nepal.

**Methods:** We performed a retrospective cross-sectional study where data were collected from medical records of adult patients (age >40 years), hospitalized with clinical diagnosis of AECOPD in a tertiary level University hospital in eastern Nepal from April 15, 2014 to October 15, 2014 were included. Data analysis was performed by using SPSS software (Version 26.0, 2020; SPSS Inc., Chicago, IL).

**Results:** Of the 256 patients with the primary diagnosis of AECOPD, mean age was 69 years and 65.63% (n=168) of hospitalizations were female population. Compared to males, 64.32 % (n=137) of active smokers were females p= 0.299, 76.19% (n=32) of diabetics were females p= 0.155, 72.86% (n=51) of hypertensive were females, p= 0.143, 50% (n= 6) of underlying Atrial fibrillation were in females p= 0.350, 57.50% (n= 23) of anemics were females p= 0.278, 100% (n= 3) of asthmatics were females p= 0.553, 44.44% (n= 8) of Pulmonary tuberculosis were in females p= 0.070, and 78.76% (n= 89) of indoor air pollution exposure was in females p <0.001.

**Conclusion:** Females have higher association to indoor air pollution exposure compared to male and this association was found to be statistically significant. The higher incidence of AECOPD hospitalization in females can be explained by these findings. We need larger studies to validate these findings.

## Introduction

Chronic Obstructive Pulmonary Disease (COPD) is defined as a progressive and incompletely reversible obstructive lung disease that is characterized by persistent respiratory symptoms and airflow limitations. AECOPD is the acute exacerbation of COPD with clinical symptoms of increased Shortness of breath or cough with or without sputum production in patients with COPD. COPD is pathologically characterized by airway and/or alveolar abnormalities usually caused by significant exposure to noxious particles or gases(1). COPD is the most common chronic respiratory condition in Nepal(2). COPD can lead to pulmonary artery hypertension resulting from the damage of structure and function of the lung. This results in right ventricular enlargement and with time may lead to right heart failure(3). Nepal is an agricultural country where females work mostly indoor cooking food with biomass fuel like firewood and cow-dung. Many of these females also smoke tobacco. According to the Nepal Demographic and Health Survey 2016, 27% of the Nepalese males and 6% of females smoked tobacco, 66% of households used solid fuel for cooking, and 31% of households were exposed to second-hand smoke or indoor air pollution (IAP) so our study also highlights the gender wise difference in prevalence of AECOPD (4).We reported the differences of comorbidities and risks between male and female gender in patients hospitalized with AECOPD in the Eastern part of Nepal.

### Pathogenesis

The pathogenesis of AECOPD involves long standing alveolar hypoxia and endothelial dysfunction by noxious particles or gases. This induces pulmonary vascular remodeling. With increasing pulmonary vascular remodeling, there is increased pulmonary vascular resistance causing Pulmonary Artery Hypertension (PAH) (5) (6). Some studies reported that the reduced lung function which is a characteristic of COPD can be a risk factor for development of DM and other co-morbidities (7, 8). Moreover, the inflammatory markers like TNF-α, IL-6, and CRP, are elevated in AECOPD (9, 10). However, the role of female gender in the development of AECOPD is not well understood.

### Objective

To explore the gender-based difference in risks and comorbidities of hospitalizations with AECOPD in Eastern-Nepal.

## Materials and Methods

This is a retrospective observational cross-sectional study where data were collected from medical records of patients admitted with clinical diagnosis of COPD in B. P. Koirala Institute of Health Sciences (BPKIHS), a tertiary level University hospital in eastern Nepal from April 15, 2014 to October 15, 2014. Our inclusion criteria:

1. Age group of more than 40 years.
2. Patients hospitalized with primary diagnosis of AECOPD

### Exclusion criteria

1. Patients with underlying structural heart disease, Cardiomyopathy, Congenital heart disease or left sided heart disease in the present or past
2. Patients with other chronic or terminal condition like cancer, transplant or HIV.

### Data items and Definitions

Diagnosis of COPD was made using GOLD criteria which includes the presence of a post-bronchodilator FEV1/FVC < 0.70 along with appropriate symptoms like dyspnea, chronic cough or sputum production, and/or history of significant exposures to noxious stimuli like cigarette smoke or household smoke from biomass fuel. Acute exacerbation of COPD (AECOPD) was diagnosed clinically based on the symptoms of increased chronic cough with or without sputum production, progressive dyspnea and progressive limitation of activity with significant exposure to noxious particles like tobacco smoke, occupational dust or household air pollution (biomass fuel smoke) or in patients with past medical history of AECOPD. Exclusion of other possible explanations and diagnosis for increased cough, dyspnea or limitation of activity example Congestive heart failure, Bronchiectasis, Interstitial Lung Disease was done with the help of Clinical history, ECG, ABG, Spirometry, Echocardiogram, Chest X-ray and Lab parameters.

### Statistical analysis and Ethical Clearance

Ethical clearance was obtained from the Institutional Review Board (IRB) of University Hospital BPKIHS. The data collection was done by two authors independently from the medical record of BPKIHS. We made a proforma and excel sheet to input all the data collected in various headings. Data analysis was performed by using SPSS software (Version 26.0, 2020; SPSS Inc., Chicago, IL). Chi square test was performed to see the association and a 2-tailed p value of <0.05 indicated statistical significance. We did not perform Univariate and multivariate regression analysis due to limited data collected during the study.

## Results

We included 256 patients with clinical diagnosis of Acute exacerbation of COPD patients in our study. The mean age of our patients is 69 years. We have illustrated the bar graph of prevalence of co-morbidities in patients hospitalized with AECOPD during the time frame in figure 1.

**Figure 1:**
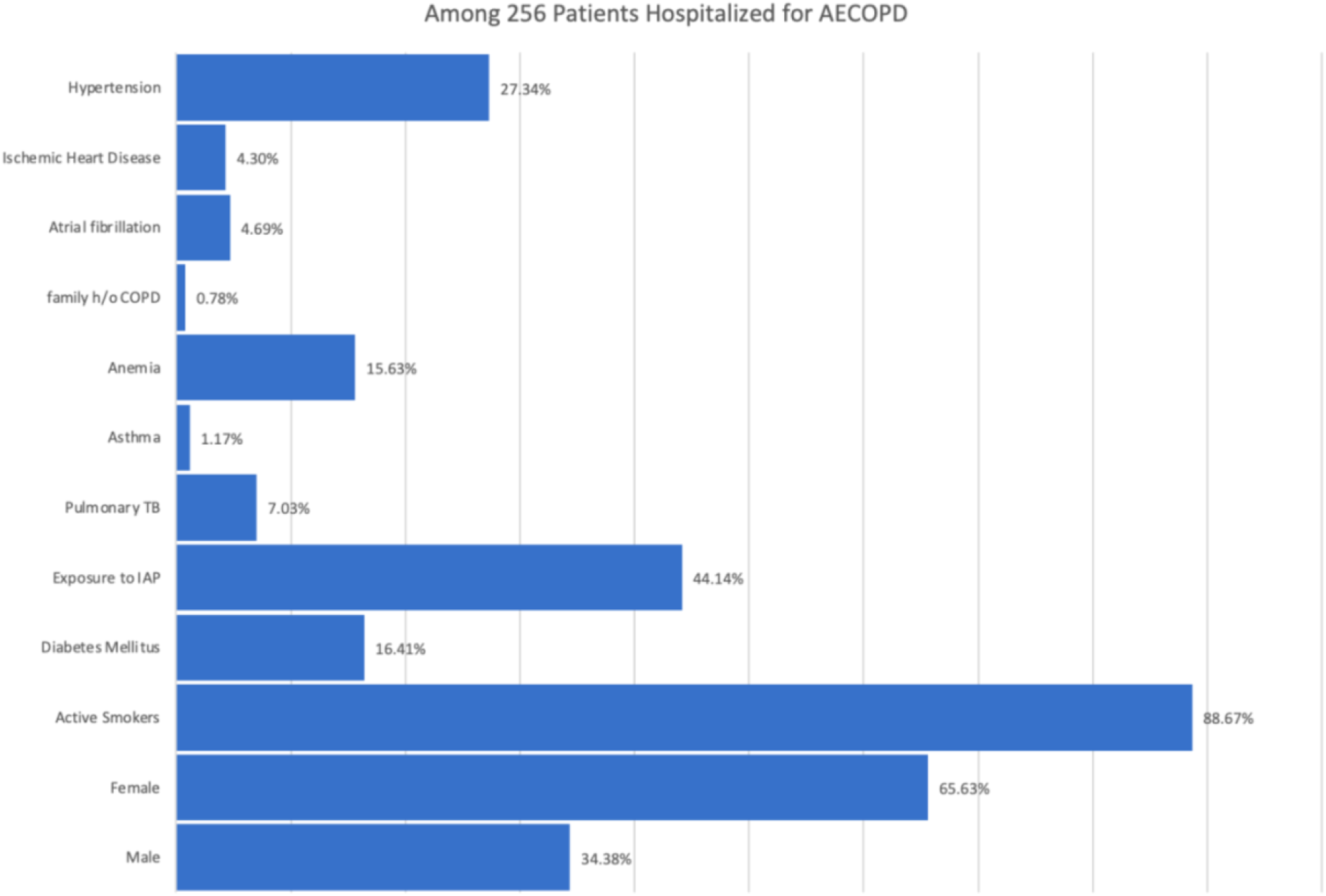
Prevalence of Comorbidities in Patient Hospitalized with AECOPD IAP= Indoor air pollution TB: Tuberculosis

Of the 256 patients with the primary diagnosis of AECOPD, mean age was 69 years, 65.63% (n=168) were female, 34.38% (n=88) were males, 88.67% (n=227) were active smokers, 16.41% (n=42) were diabetic, 44.14% (n=113) were exposed to household air pollution, 7.03% (n=18) had pulmonary tuberculosis, 1.17% (n=3) had underlying asthma, 15.63% (n=40) were anemic, 0.78% (n=2) had history of COPD in family, 4.69% (n=12) had underlying Atrial fibrillation, 4.30% (n=11) had ischemic heart disease, and 27.34% (n=70) were hypertensive. Compared to males, 64.32 % (n=137) of active smokers were females p= 0.299, 76.19% (n=32) of diabetics were females p= 0.155, 72.86% (n=51) of hypertensive were females, p= 0.143, 50% (n= 6) of underlying Atrial fibrillation occurred in females p= 0.350, 57.50% (n= 23) of underlying anemia occurred in females p= 0.278, 100% (n= 3) of asthmatics were females p= 0.553, 44.44% (n= 8) of Pulmonary tuberculosis occurred in females p= 0.070), and 78.76% (n= 89) of indoor air pollution exposure occurred in females p <0.001).

## Discussion

Our study shows the higher incidence of hospitalization for AECOPD in females of Eastern-Nepal and this can be explained by high exposure to IAP/biomass fuel smoke as well as high prevalence of smoking in females, compared to male. The combined effect of active and passive smoking along with indoor air pollution may also have contributed to the higher association of female gender with development of COPD. This is plausible from the socio-cultural aspect of Nepal where females mostly work indoors and houses still use biomass for fuel which generates a lot of indoor smoke (2). The study by Jain et al also showed higher association of IAP and COPD with female gender having a prevalence of 89% (11). A meta-analysis by Hu G et al also showed the additive effect of smoking with IAP to cause significant damage to the lungs leading to the development of COPD and its complications (12). This explains the increased incidence of AECOPD in female gender in Eastern-Nepal due to higher IAP exposure and smoking habits in female compared to male.

There is paucity of data on hospitalized patient with AECOPD and the gender based differences in the prevalence of comorbidities (13). Various studies have reported the prevalence of diabetes in patients with COPD from 1.6-16%(14). The study by Curkendall et al found the prevalence of DM as 14.5 % and HTN as 40.6% in patients with COPD (15). The use of chronic steroids in patients with COPD can be associated with the increased risk of having DM in these patients. A study by Mahishale et al found the prevalence of DM and HTN in the 2432 COPD subjects was 25.94% and 37.25% respectively which is higher than reported by our study. However, our study differs from other studies as our population includes hospitalized patients with AECOPD. His study reports that severe COPD(GOLD stage 3 or 4) was associated with a higher risk of DM (odds ratio OR 1.6, 95% CI 1.2–2) and HTN (OR 1.6, 95% CI 1.4–1.9)(16). This is supported by the study by Mannino et al which reports that after adjusting for age, sex, race, smoking, body mass index and education, subjects with GOLD stage 3 or 4 COPD had a higher prevalence of DM (odds ratio (OR) 1.5, 95% confidence interval (CI) 1.1–1.9) and hypertension (OR 1.6, 95% CI 1.3–1.9). Patient with more co-morbidities like diabetes, hypertension, and cardiovascular disease were associated with a higher risk of hospitalization and mortality especially in patients with severe COPD (17).

Our study has several limitations including smaller sample size, lack of proper adjustment of all the potential confounder, and bias in the study design. Moreover, since it was done in eastern-Nepal in a single hospital setting, it may not be generalizable to the overall population. The retrospective chart review is prone to a lot of biases including selection bias, measurement bias, and observer-expectancy bias. Many important information may have been missed during the documentation. However, all efforts are made to decrease the bias, including complete chart review and second review of charts. We did not comment on in-hospital mortality or readmissions in our study due to the cross-sectional nature of our study and lack of in-hospital mortality as one of our variables. Our study highlights that we need larger studies to comment on the gender-based differences in patients with AECOPD.

## Conclusions

Our study highlights females of Eastern-Nepal have higher association to indoor air pollution exposure compared to males and this association was found to be statistically significant. The higher incidence of AECOPD hospitalization in females in this population can be explained by these findings. We need larger studies to explain the complex association between female gender and AECOPD.

## Data Availability

Please email at for the data.

## Conflict of Interest

We have no any conflict of interest for this study and this study is not funded by any party.

## ACKNOWLEDMENTS

None

## Notes

### Competing Interest Statement

The authors have declared no competing interest.

### Clinical Trial

not registered

### Author Declarations

Ethical clearance from Institutional Review Committee, B. P. Koirala Institute of Health Sciences Dharan, Nepal has been granted on April 9, 2015 Our study was approved from the IRC. The consent was waived based on the nature and quality of the study as follows. The study is a retrospective chart review of hospitalizations for COPD exacerbation. The research fits criteria of consent waiver from IRC of BPKIHS due to: 1. The research involves the collection of limited de-identified data from medical charts of BPKIHS retrospectively AND 2. The data is recorded by the investigator in an anonymous manner such that subjects cannot be identified directly or through identifiers linked to the subject. Approved by : Head of Department of Internal Medicine and Chair of Pulmonology, Critical Care and Sleep medicine: Prof. Dr. Narendra Bhatta

